# Multidrug-Resistant Bacteria Causing Post-Caesarian Section Surgical Site Infection at Regional Referral Hospitals in Dar es Salaam, Tanzania

**DOI:** 10.1101/2025.08.01.25332674

**Authors:** Biswaro Malima, Mtebe Majigo, Damas Chipaga, Joel Manyahi, Agricola Joachim, Said Aboud

**Affiliations:** Department of Microbiology and Immunology, Muhimbili University of Health and Allied Sciences, Dar es Salaam, Tanzania

**Keywords:** Caesarean Section, Multidrug Resistance, Surgical Site Infection, Infection Prevention, Control

## Abstract

**Background:** Surgical site infections (SSI) are the most common nosocomial infection among surgical patients. When infection is associated with multidrug-resistant bacteria, it leads to poor prognosis and increased morbidity, mortality, hospital stay, and cost of treatment. This study determined the etiological agents of SSI and their antimicrobial susceptibility pattern in post-caesarian section women at Regional Referral Hospitals in Dar es Salaam, Tanzania.

**Methods:** We conducted a cross-sectional study between June 2022 and January 2023 by enrolling 175 patients with signs and symptoms of SSI who provided informed consent. A structured questionnaire was used to collect demographic and clinical information. Under aseptic techniques, pus swabs were collected for aerobic bacterial culture. Isolates were identified using Analytical Profile Index-20E. Antimicrobial susceptibility tests were performed using Kirby-Bauer disc diffusion and interpreted per clinical laboratory standard institutes 2022 guidelines. We performed descriptive statistics using STARTA version 15 and summarized continuous variables as mean and standard deviation and categorical variables as proportions.

**Results:** Multiple 185 pathogens were isolated, the majority, 129(69.7%) being gram-negative. The predominant bacteria was *Klebsiella pneumoniae*, 58(29.9%), followed by *Staphylococcus aureus*, 55(28.3%). The extended-spectrum beta-lactamase production was observed in 35(38.5%) Enterobacterales. Of 55 *Staphylococcus aureus*, 29(52.7%) were methicillin-resistant and 19(34.5%) inducible clindamycin resistant. The overall proportion of multidrug-resistant bacteria was 145(78.8%), observed more in *Klebsiella* spp (91.7%), *Staphylococcus aureus* (85.5%) and *Pseudomonas aeruginosa* (46.7%). Most gram-negative bacteria, 72(55.8%), were resistant to more than four antibiotic classes.

**Conclusion:** Various pathogens, predominantly Gram-negative bacteria, caused prost-caesarian section SSI. More than three-quarters of the pathogens were multidrug-resistant, including commonly prescribed antibiotics. The findings call for strengthening infection prevention and antimicrobial stewardship interventions.

## INTRODUCTION

Surgical site infections (SSIs) are common complications of caesarean section that contribute significantly to mortality, prolonged hospital stays, and increased healthcare costs (1). Worldwide, the incidence of SSI ranges from 1.2 to 23.6 per 100 surgical procedures (2,3). They are the most common healthcare-associated infections in obstetrics and represent a considerable burden for healthcare systems, especially in low- and middle-income countries (LMICs) (2). Studies in Tanzania reported SSI ranging from 19.4% to 36.5% (4-7)

The organisms that cause SSI may be derived from the endogenous environment, surgical instruments, or theatre environments (8). Several bacterial pathogens have been found as the causative agents of SSI with varying antimicrobial susceptibility patterns. Studies conducted in Africa report high resistance to commonly used antibiotics (9-11). In addition, most pathogens were resistant to multiple drugs, with the proportion of multi-drug resistant (MDR) bacteria ranging from 63%-86% (12-19).

The tremendous change in the community’s economic and social lifestyle facilitates the increase in cesarean sections and, subsequently, the occurrence of SSI (20). Increasing antimicrobial resistance with time requires a periodic study to generate data regarding the magnitude and pattern of antimicrobial susceptibility in SSI. We conducted the study at regional referral hospitals with high antibiotic use levels. The study findings inform the best practices for prophylaxis and managing SSI.

## MATERIALS AND METHODS

### Study design and setting

We conducted a cross-sectional study from June 2022 to January 2023 among women post-cesarean sections at Amana, Mwananyamala, and Temeke Regional Referral Hospitals in Dar es Salaam, Tanzania. The three hospitals were selected due to the high volume of clients undergoing cesarian sections, approximately 780 monthly. Amana Regional Referral Hospital is located in the Ilala district, has a 382-bed capacity, and attends approximately 280 post-caesarian clients monthly. Temeke Regional Referral Hospital is located in Temeke district, has a capacity of 324 beds, and attends approximately 260 clients post-cesarian sections monthly. Mwananyamala Regional Referral Hospital is situated in Kinondoni district; it has a 317-bed capacity and attends approximately 240 post-cesarian sections women per month.

### Patient enrollment and data collection

Post-cesarean sections, women with clinical features of SSI were eligible to be included in the study. We enrolled participants from inpatients and outpatients who provided written informed consent. Outpatients were assessed, and those who met the criteria were recruited at the time of stitches removal. The study used a structured questionnaire to collect participants’ socio-demographic and clinical information. Under the aseptic technique, two pus swabs were collected from the infected surgical wound and immersed in an Amies transport medium. The specimens were transported to the Muhimbili University of Health and Allied Sciences (MUHAS) bacteriology Laboratory for processing within two hours of collection.

### Bacterial Isolation and Identification

Gram staining was performed from one of the two swabs, while the other was inoculated on the blood agar (Oxoid, UK) and MacConkey agar (Oxoid, UK) plates. The plates were incubated aerobically at 37^°^C for 24 hours. If the Gram staining showed the presence of gram-positive cocci in chains, a bacitracin disc was added to the blood agar plate. Bacterial isolates were characterized using growth pattern, colony morphology, hemolysis pattern on blood agar plate, gram stain, and conventional biochemical tests based on the gram reaction (21). A catalase test was performed for gram-positive cocci to differentiate *Staphylococcus* and *Streptococci* species. The DNase test confirmed *Staphylococcus aureus*, while bile esculin, bacitracin, and optochin disc were used to identify *Streptococcus* species. Gram-negative rods were identified through biochemical tests like oxidase, urease, citrate, indole, motility, hydrogen sulfide, and Kligler iron agar. We used analytical profile index-20 (AP20E) to identify gram-negative isolates with inconclusive conventional biochemical tests.

### Antimicrobial susceptibility testing

We performed antimicrobial susceptibility testing (AST) using the Kirby Bauer disc diffusion method. The AST results were interpreted according to the Clinical and Laboratory Standards Institute guidelines 2022. Briefly, test and control organisms were suspended in normal saline equivalent to McFarland 0.5 standard and inoculated on Mueller-Hinton agar (Oxoid, UK). Appropriate six discs were placed into a 90mm plate and incubated aerobically at 37°C for 16 to 18 hours. All antibiotic discs used for AST were made from Oxoid, UK.

We used penicillin-G (10IU), gentamycin (10μg), sulfamethoxazole/trimethoprim (1.25/23.75 μg), clindamycin (2μg), erythromycin (15μg), chloramphenicol (30μg), tetracycline (30μg), doxycycline (5μg) and ciprofloxacin (5μg) for gram-positive bacteria. In addition, cefoxitin (30μg) was included for *Staphylococcus* species. Enterobacteriaceae were tested with ampicillin (10μg), amoxicillin/clavulanic acid (20/10μg), ciprofloxacin (5μg), gentamicin (10μg), sulfamethoxazole/trimethoprim (1.25/23.75μg), chloramphenicol (30μg) ceftriaxone (30μg), ceftazidime (30μg), amikacin (30μg), meropenem (10μg), cefepime (30μg), tetracycline (30μg) and tazobactam (10μg). *Pseudomonas aeruginosa* were tested using ciprofloxacin (5μg), gentamicin (10μg), ceftazidime (30μg), amikacin (30μg), meropenem (10μg), cefepime (30μg) and tazobactam (10μg). *Acinetobacter baumanii* were tested using sulfamethoxazole/trimethoprim (1.25/23.75μg), ciprofloxacin (5μg), gentamicin (10μg), chloramphenicol (30μg), ceftriaxone (30μg), ceftazidime (30μg), amikacin (30μg), meropenem (10μg), cefepime (30μg), tetracycline (30μg) and tazobactam (10μg). The AST result was classified as susceptible, intermediate, and resistant.

### Detection of antimicrobial resistance phenotypes

Methicillin-resistant *Staphylococcus aureus* (MRSA) was determined using a cefoxitin disc (30 μg) during AST. A zone of inhibition equal to or less than 21mm was considered positive. Inducible clindamycin resistance (ICR) was detected using erythromycin (15μg) and clindamycin (2μg) discs placed 15mm apart during AST. The formation of a D-shaped toward erythromycin was regarded as positive. We confirmed the production of extended-spectrum beta-lactamase (ESBL) among gram-negative isolates using a combination method. The ceftriaxone and ceftazidime discs were used alone and in combination with clavulanic acid. An increase of 5mm or more in the zone of inhibition around the combined discs confirmed ESBL production.

### Quality control

Aseptic techniques were observed during specimen collection, media preparation, and bacterial culture. Standard quality control strains were used to validate the performance of the biochemical reagents/test kits and monitor the accuracy and precision of susceptibility testing procedures and antibiotic discs. We used *Escherichia coli* ATCC 25922 for gram-negative bacteria, *Klebsiella pneumoniae* ATCC 700603 for ESBL-positive, *Staphylococcus aureus* ATCC 25923 for gram-positive bacteria, and *Staphylococcus aureus* ATCC 29213 for MRSA positive.

### Data analysis

We used STATA version 15.1 software for statistical analysis. Continuous variables were summarized as the median and interquartile range (IQR), whereas proportions were used to describe categorical variables. Data were analyzed and processed through editing, coding, tabulation, and pictorial presentation. Also, cross-tabulation was done to determine frequencies regarding the respective research objectives, and a p-value < 0.05 was considered significant. MDR was revealed if an isolate was resistant to one or more agents in at least three classes of antibiotics.

## RESULTS

### Socio-demographic and clinical characteristics

We recruited 175 patients clinically diagnosed with post-caesarian section SSI with a mean age of 28.8±5.2 years and parity of 2±1 children. Most participants, 117(66.9%), had attained primary education and 133(76.0%) were self-employed. The majority of participants, 160(91.4%), were HIV-negative tests and 164(93.4%) reached term gestation age, and 168(96.0%) were given pre-operative prophylaxis. Among those who received prophylaxis, 168(100.0%) received metronidazole, 140(83.3%) ceftriaxone, and 28(16.7%) ampicillin. About 167(95.5%) of patients had an emergency cesarean section, and 71(40.6%) had a history of antimicrobial use within the past three months before the cesarean section. Among all patients, 56(32.0%) had a history of admission within the past three months before the caesarian section, with the common type incision being 130 (74.3%) Pfannenstiel. (Table 1)

**Table 1:** Socio demographic and clinical characteristics of study participants (N=l75)

| **Variable** |  | **Frequency** | **Percent (%)** |
| --- | --- | --- | --- |
| **Age in years** | Mean (SD) | 28.8(5.2) |  |
|  | <26 | 45 | 25.7 |
|  | 26–35 | 113 | 64.6 |
|  | >36 | 17 | 9.7 |
| **Parity** | Mean (SD) | 2(1) |  |
|  | <2 | 90 | 51.4 |
|  | 2 | 55 | 31.4 |
|  | >3 | 30 | 17.2 |
| **Study site** | Amana | 72 | 41.1 |
|  | Mwananyamala | 22 | 12.6 |
|  | Temeke | 81 | 46.3 |
| **Gestation age** | Preterm | 11 | 6.3 |
|  | Term | 164 | 93.7 |
| **Marital status** | Single | 12 | 6.9 |
|  | Married | 160 | 91.4 |
|  | Unmarried | 3 | 1.7 |
| **Education level** | Primary | 117 | 66.9 |
|  | Secondary | 34 | 19.4 |
|  | College/University | 24 | 13.7 |
| **Occupation** | Employed | 9 | 5.1 |
|  | Self-employed | 133 | 76.0 |
|  | Unemployed | 33 | 18.9 |
| **HIV status** | Positive | 15 | 8.6 |
|  | Negative | 160 | 91.4 |
| **Antibiotic prophylaxis** | Yes | 168 | 96.0 |
| **Used Antibiotics** | Metronidazole | 168 | 100.0 |
|  | Ceftriaxone | 140 | 83.3 |
|  | Ampicillin | 28 | 16.7 |
| **Type of caesarian section** | Elective | 8 | 4.6 |
|  | Emergency | 167 | 95.4 |
| **Antibiotics use with 3months** | Yes | 71 | 40.6 |
| **Admission in three months** | Yes | 56 | 32.0 |
| **Type of Incision** | Pfannenstiel | 130 | 74.3 |
|  | Sub-umbilical midline incision | 45 | 25.7 |

HIV-Human immunodeficiency virus

### Bacterial Etiology of SSI

Out of 175 samples, 164(93.7%) were culture positive, 143/164(87.2%) of the culture-positive were pure isolates, while 21/164(12.8%) had two significant isolates. Gram-negative pathogens were 129/185(69.7%). Overall predominant pathogen was *Klebsiella pneumoniae*, 58(31.4%), followed by *Staphylococcus aureus*, 55(29.7%), and *P. aeruginosa* 30(16.2%) (Figure 1)

**Fig. 1.**
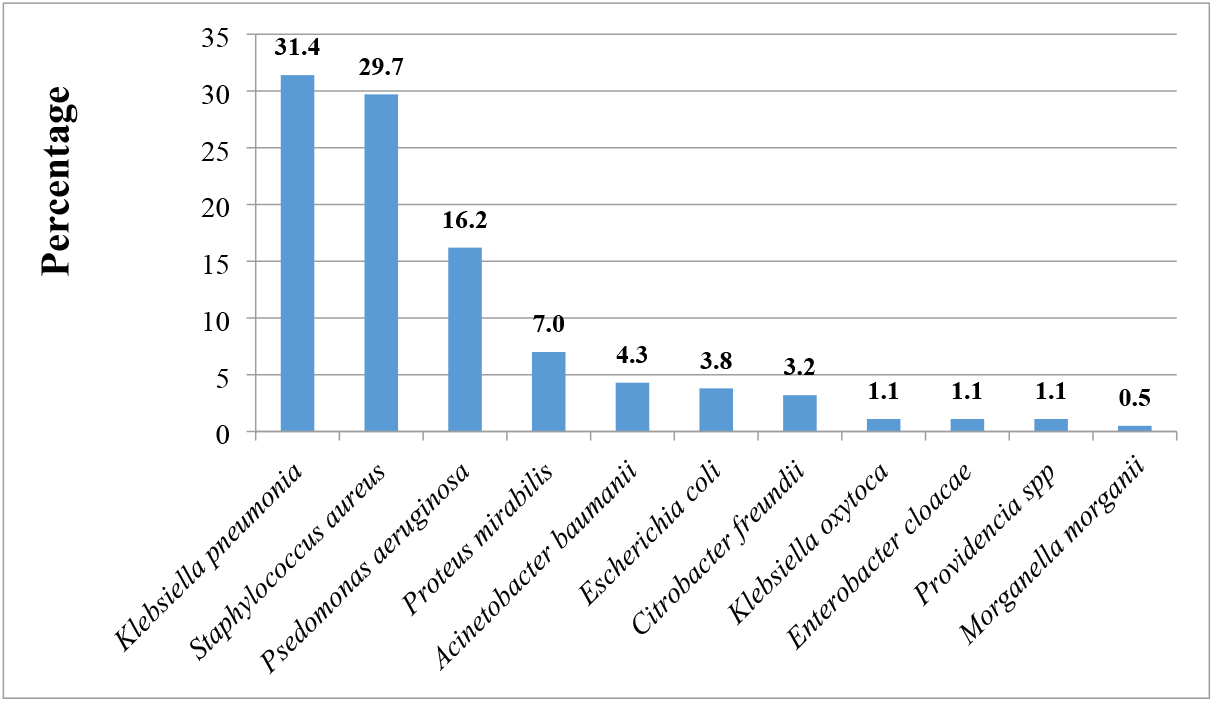
Percent distribution of bacteria isolated in post-caesarian section surgical site infection.

### Antimicrobial resistance pattern

*Staphylococcus aureus* showed high resistance to penicillin (87.3%), followed by erythromycin (74.6%) and moderate resistance to trimethoprim-sulfamethoxazole (58.2%), clindamycin (41.8%), chloramphenicol (36.4%), and ciprofloxacin (30.9%). Low resistance for *Staphylococcus aureus* was observed against tetracycline (27.3%), gentamycin (20.0%), and doxycycline (16.4%). Gram-negative bacteria showed high resistance to ampicillin (97.8%), cefotaxime (80.8%), and ceftriaxone (80.8%). Moderate resistance for Gram-negative was observed against chloramphenicol (61.6%), ceftazidime (57.4%), amoxicillin-clavulanic acid (54.9%), and ciprofloxacin (52.2%). The least resistance was observed against amikacin (3.1%). The most predominant gram-negative isolate, *Klebsiella* species, showed high resistance against ampicillin (100%), ceftriaxone (96.7%), trimethoprim-sulfamethoxazole (81.7%), ceftazidime (75.0%), amoxicillin-clavulanic acid (70.7%), chloramphenicol (63.3%) and cefepime (63.3%). Moderate resistance for *Klebsiella* species was found against tetracycline (58.3%) and ciprofloxacin (46.7%), while low resistance was observed against tazobactam (28.3%), gentamycin (28.3%), meropenem (25.0%), and amikacin (3.3%). (Table 2)

**Table 2:** Proportion of antimicrobial resistance of bacterial isolates among post-caesarian section women with S51 at Regional Referral.

| Bacterial isolate | Percentage of Resistant Pathogens to Antimicrobial Agents (%) |  |  |  |  |  |  |  |  |  |  |  |  |  |  |  |  |  |  |
| --- | --- | --- | --- | --- | --- | --- | --- | --- | --- | --- | --- | --- | --- | --- | --- | --- | --- | --- | --- |
|  | MEM | AMP | CIN | AK | FEP | TZP | CRO | CTX | CAZ | AUG | CIP | SXT | C | TE | FOX | DO | CD | E | PEN |
| *S aureus* (N=55) | NA | NA | 20.0 | NA | NA | NA | NA | NA | NA | NA | 30.9 | 58.2 | 36.4 | 27.3 | 52.7 | 16.4 | 41.8 | 74.6 | 87.3 |
| *Enterococcus faecalis* (N=1) | NA | NA | 0 | NA | NA | NA | NA | NA | NA | NA | 0 | 0 | 0 | 0 | NA | 0 | 100 | 0 | 0 |
| *E coli* (N=7) | 0 | 100 | 14.3 | 0 | 14.3 | 14.3 | 71.4 | 71.4 | 42.9 | 42.9 | 42.9 | 100 | 71.4 | 71.4 | NA | NA | NA | NA | NA |
| *Klebsiella spp* (N=60) | 25.0 | 100 | 28.3 | 3.3 | 63.3 | 28.3 | 96.7 | 96.7 | 75.0 | 70.7 | 46.7 | 81.7 | 63.3 | 58.3 | NA | NA | NA | NA | NA |
| *P. aeruginosa* (N=30) | 40.0 | NA | 43.3 | 6.7 | 70.0 | 46.7 | NA | NA | 53.3 | NA | 50.0 | NA | NA | NA | NA | NA | NA | NA | NA |
| *P. mirabilis* (N=13) | 0 | 84.6 | 7.7 | 0 | 7.7 | 0 | 23.1 | 23.1 | 7.7 | 15.4 | 15.4 | 53.8 | 30.8 | 46.2 | NA | NA | NA | NA | NA |
| *A. baumannii* (N=8) | 25 | NA | 37.5 | 0 | 75.0 | 37.5 | 100 | 100 | 87.5 | NA | 50.0 | 100 | 87.5 | 75.0 | NA | NA | NA | NA | NA |
| *Other Enterobactrales* (N=11) | 9.1 | 100 | 9.1 | 0 | 45.5 | 27.3 | 54.5 | 54.5 | 18.2 | 27.3 | 36.4 | 45.5 | 63.6 | 63.6 | NA | NA | NA | NA | NA |
| Total | 23.3 | 97.8 | 25.4 | 3.1 | 55.8 | 29.5 | 80.8 | 80.8 | 57.4 | 54.9 | 39.5 | 69.7 | 52.2 | 47.7 | 52.7 | 16.5 | 42.9 | 74.6 | 87.3 |

MEM-meropenem (10µg), AMP-ampicillin (10µg), CN-gentamycin (10µg), AK-amikacin (30µg), FEP-cefepime (30µg), TZP-tazobactam (10µg), CRO-ceftriaxone (30µg), CTX-cefotaxime (30µg), CAZ-ceftazidime (30µg), AUG-augmentin (30µg), CIP-ciprofloxacin (5µg), SXT-trimethoprim sulfamethoxazole (1.25/23.7µg), C-chloramphenicol (30µg), TE-tetracycline (30µg), PEN-penicillin (10IU), FOX-cefoxitin (30µg), DO-doxycycline (5µg), CD-clindamycin (2µg), E-erythromycin (15µg), NA- Not Applicable.

### Magnitude of multi-drug resistant bacteria

The overall proportion of MDR of isolated bacteria was 78.9%. The proportion of MDR for *S. aureus, Klebsiella* spp, *P. mirabilis*, and *P. aeruginosa* was 85.5%, 91.7%, 53.8%, and 46.7%, respectively. All isolates of *A. baumannii* and *Escherichia coli* were MDR. Among other Enterobacterioles (*C freudii, E. cloacae, Providencia* spp, *and M. morganii*), 63.7% were MDR strains. About 68.5% of isolates were resistant to four or more antimicrobial classes, while 80.0% of the *Klebsiella* spp were resistant to ≥5 classes of antimicrobial agents. We also noted that 42.9% of the *Escherichia coli* isolates were resistant to ≥6 classes of antimicrobial agents, and 32.7% of the *Staphylococcus aureus* isolates were resistant to at least six classes of antimicrobial agents. (Table 3).

**Table 3.** The proportion or multidrug resistance in bacteria isolated from women with SSI.

| Bacteria isolates | Percentage of Resistance |  |  |  |  |  |  |  |
| --- | --- | --- | --- | --- | --- | --- | --- | --- |
|  | ≤ % R2 | % R3 | % R4 | % R5 | % R6 | % R7 | % R8 | % MDR |
| *S. aureus* (N= 55) | 14.5 | 21.8 | 16.4 | 14.6 | 12.7 | 14.6 | 5.5 | 85.5 |
| *E. coli* (N=7) | - | - | 14.3 | 42.9 | 42.9 | - | - | 100.0 |
| *Klebsiella* spp (N=60) | 8.3 | - | 11.7 | 26.7 | 20.0 | 15.0 | 18.3 | 91.7 |
| *P. aeruginosa* (N=30) | 53.3 | 6.7 | 16.7 | 23.3 | - | - | - | 46.7 |
| *P. mirabilis* (N=13) | 46.2 | 23.1 | 23.1 | 7.7 | - | - | - | 53.8 |
| *A. baumannii* (N=8) | - | 12.5 | 25.0 | 25.0 | 25.0 | - | 12.5 | 100 |
| *Other Enterobacterales* (N=11) | 36.3 | 9.1 | 9.1 | 9.1 | 18.2 | 9.1 | 9.1 | 63.7 |
| Total | 21.1 | 10.4 | 15.7 | 20.5 | 14.3 | 9.8 | 8.2 | 78.9 |

R-Antibiotic Resistance Class, *Klebsiella* Spp (*K. pneumoniae* and *K. oxytoca*), Other Enterobacterales ( *C. freundii*, *E. cloacae*, *Providencia* spp and *M. morganii*)

### Proportion of MRSA, ICR, and ESBL

Of the 55 *S. aureus* isolates, 52.7% (29/55) were MRSA strains, and the proportion of ICR was 34.5% (19/55). Among the 91 gram-negative bacterial isolates, 35(38.5%) were ESBL-producing strains. (Figure 2)

**Fig. 2.**
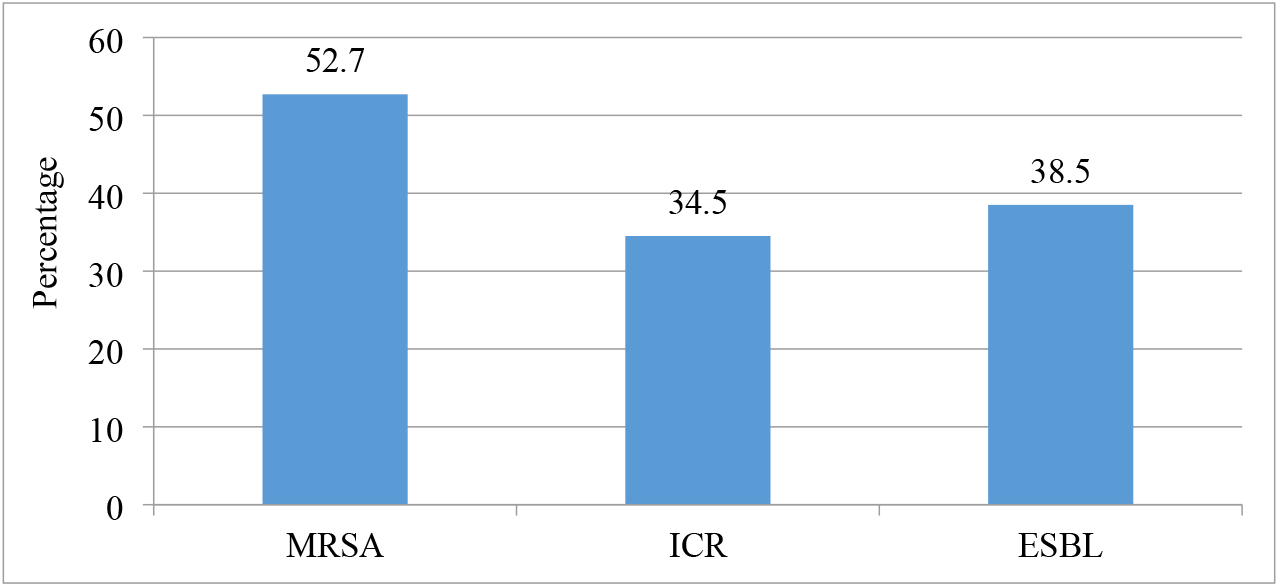
Proportion of MRSA, CIR, and ESBL among the isolated bacteria. MRSA-Methicillin Resistance *Staphylococcus aureus*, ICR-Induced Clindamycin Resistance, and ESBL Extended-spectrum beta-lactamase

## DISCUSSION

We found that gram-negative bacteria were the dominant cause of SSIs. Most pathogens were resistant to ceftriaxone and ampicillin, the commonly prescribed antimicrobial agents. In addition, more than three-quarters (78.9%) of pathogens causing SSI were MDR. The dominance of gram-negative bacteria in SSIs presents significant clinical challenges due to their inherent resistance mechanisms and ability to cause severe infections. The MDR pathogens limit treatment options and, as a result, prolong hospital stays, increase healthcare costs, and heighten the risk of adverse outcomes. The findings highlight the need for surveillance, antimicrobial stewardship programs, and infection control measures to prevent further emergence and transmission of MDR.

The predominance of gram-negative bacteria in SSIs in the current studies concurs with other studies in the country and other parts of the world (4,8,22). The study found the predominant isolate was *K. pneumoniae*, similar to studies conducted in India, Uganda, and Ethiopia (8,12,14,23). The similarity might be due to the proximity of the incision site to where most of these endogenous microbiotas reside. *Klebsiella* species have been reported as common contaminants in operating room air and fomites, including hospital medical equipment (24). Contrary to our findings, other studies reported *S. aureus* as the dominant pathogen in post-caesarian section SSI (10,11,17,19), and a study conducted at MNH, Tanzania, reported *P. aeruginosa* as the most common isolate (15). The difference between our findings and the study at MNH might be due to different study populations; the previous study included all patients from all departments.

*Klebsiella* spp, the most predominant pathogen in the current study, was highly resistant to commonly used antibiotics like ampicillin, ceftriaxone, trimethoprim-sulfamethoxazole, and amoxicillin-clavulanic acid. Similarly, *S. aureus*, the second predominant pathogen, was highly resistant to penicillin. A similar finding was observed in another study (25). The level of AMR among gram-negative bacteria (53-100%) to antibiotics for prophylaxis and treatment (ceftriaxone, amoxicillin-clavulanic acid, cotrimoxazole, ampicillin, and cloxacillin) are in keeping with reports from Tanzania and Uganda (4,14,15). The observed resistance rate to these antibiotics may be due to their frequent use empirically. Also, cotrimoxazole is widely used for prophylaxis against opportunistic infections in people living with HIV. Resistance to third-generation cephalosporin, like ceftriaxone (80.8%), was higher among gram-negative bacteria. Most *E. coli* and *K. pneumoniae* were ESBL-producing strains, as reported in other Uganda and Ethiopia studies (10, 14). Overusing ceftriaxone as a broad-spectrum antibiotic for pre-operative prophylaxis and empirical treatment of other bacterial infections may contribute to the emergency of resistance (26). However, we have observed very low resistance to amikacin, which might be due to limited exposure to this antibiotic.

A significant proportion of the *A. baumannii* demonstrated high levels of resistance against a wide array of antimicrobial agents, ranging from 25% to 100%, similar to a previous study conducted in the same setting (15). The high resistance profile indicates a concerning trend in the efficacy of common antibiotics against this particular pathogen. Such widespread resistance poses serious challenges in the clinical management of infections caused by *A. baumannii*. The ability of this pathogen to withstand the effects of multiple antimicrobial agents suggests the presence of various resistance mechanisms.

We observed more than three-quarters (78.9%) of bacterial pathogens being MDR, which was slightly low compared to 82.92% reported in Ethiopia (25) and 86.0% in Uganda (8). On the other hand, the proportion of MDR pathogens found in the current study was much higher than in the previous study (63%) conducted about ten years ago (15). About 68.6% of isolates had developed resistance to four or more antimicrobial classes, and 55.8% of the gram-negative bacteria were resistant to ≥ 5 antibiotic classes. We noted that all *E. coli, A. baumanii, CONS*, and *E. cloacae* isolated were MDR stains. The findings on MDR add evidence to the global threat of AMR and could indicate a progressive increase in MDR with time.

The current study investigated aerobic bacteria only as a cause of post-caesarian section SSI. This limitation leads to an incomplete understanding of the overall microbial population in this infection. Future studies should include an investigation of anaerobes, which require careful handling and special environments for growth. Another limitation is that we did not follow up with patients after discharge from the hospital to evaluate the effectiveness of the prescribed antibiotics and make adjustments if needed to combat the resistant pathogens.

## CONCLUSIONS

The most common isolates involved in the post-cesarian section SSIs at the Regional Referral Hospitals in Dar es Salaam were *K. pneumoniae, S. aureus*, and *P. aeruginosa*. Most species were resistant to antibiotics commonly prescribed for prophylaxis and empirical treatment of SSI. We observed a high proportion of MDR bacteria with an increased prevalence of ESBL and MRSA. The emergence of multidrug-resistant bacteria underscores the urgent need to strengthen AMR surveillance, infection prevention and control measures, and antimicrobial stewardship programs. Routine culture and AST for post-cesarian section SSIs are essential to optimize antimicrobial use for each patient.

## Data Availability

XXX, XXX

## Abbreviation

AMR-: Antimicrobial Resistance
CS-: Caesarean Section
HAI-: Health Care-Associated Infection
IPC-: Infection Prevention and Control
ICR-: Inducible Clindamycin Resistance
MDR-: Multidrug Resistance
MUHAS-: Muhimbili University of Health and Allied Sciences
MRSA-: Methicillin Resistance *Staphylococcus aureus*
SSI-: Surgical Site Infection

## Ethical considerations

We obtained ethical approval from the MUHAS Research and Ethics Committee, Reference No. DA.282/298/01.C/MUHAS-REC-02-2022-990. The management of regional referral hospitals permitted the enrollment of participants. Before enrolling in the study, each participant provided written informed consent.

## Acknowledgments

We thank all the patients who participated in this study for their willingness and cooperation in advancing medical knowledge and improving patient care. We also appreciate the support from the staff at the hospitals involved in the study and their cooperation during the study period.

